# Multiple Burdens of Road Traffic Crashes in Pokhara, Nepal: A Patient Approach

**DOI:** 10.1101/2022.05.16.22275143

**Authors:** Sapana Bishwokarma, Chiranjivi Adhikari, Dhurba Khatri, Bikash Gauchan, Vishnu Prasad Sapkota, Chhabi Lal Ranabhat

**Affiliations:** Pokhara University, School of Health and Allied Sciences, Pokhara, 33700, Kaski, Nepal; Indian Institute of Public Health Gandhinagar (IIPHG), Gandhinagar, 382042, India; Infectious and Tropical Disease Hospital, Pokhara, Kaski, Nepal; Department of Economics, Nepal Commerce Campus, Tribhuvan University, Nepal; Department of Health Promotion and Administration, Eastern Kentucky University, USA

**Keywords:** Road traffic crash, RTC, DALYs, Cost of illness, Disability, Nepal

## Abstract

**Background:** The economic burden and cost-related evidence from primary data of road traffic crashes (RTCs) in Nepal are limited.

**Aim:** This study assessed the disability-adjusted life years (DALYs), the cost associated with RTCs, and socio-demographic and injury related factors.

**Methods:** We carried out a retrospective cross-sectional, institution-based survey of the RTC victims in the last one year to 45 days prior to the survey, in Pokhara and the surroundings in Kaski district, a mid-hilly district and the headquarter of Gandaki province of Nepal. A sample of 107 RTC victims, registered in different hospitals, police offices and health insurance offices, were retrospectively approached. A semi-structured interview schedule was used to collect data. Data were entered into EpiData before being imported, and analysed in the Statistical Package for Social Sciences (SPSS) and MS-Excel. Descriptive statistics were generated by calculating costs and DALYs, and inferential analysis was used to investigate the relationship between DALYs and independent variables.

**Results:** More than one-third (37.4%) of the 107 victims were between the ages of 25 and 44, and more than half (60.7%) were men. The cost per million person-years of RTC was around US $ 82,800. Similarly, around 1021 DALYs per million person-years are attributed to RTC, and more than 99% of them are shared by YLL. We observed the highest number of frequencies of young (p<.01), having university degree (p<.01), unemployed (p<.05), motorcyclists (p<.01), highest economic quintile (p<.01), and seriously injured (p<.01). In addition, we also observed a low positive correlation (r=0.33, p<.01) between DALYs and the victim’s indirect cost.

**Conclusion:** The cost and DALYs associated by RTC of Kaski district were around US $ 82,800 and 1021 per million person-years in 2017, respectively. Young unemployed motorcyclists were the most frequent victims. Victim’s indirect cost is increased with DALYs, suggesting a policy implication.

## 1. Introduction

A road traffic crash (RTC) is a collision or incident involving at least one moving vehicle, in which the injured may die or seek medical treatment or assistance within 30 days of the trauma (1, 2). Based on the severity of the incident, an RTC can be fatal or non-fatal, whereas the outcome intensity may result in fatal, serious, and slight injuries (3, 4). The overall definition and analysis of the severity of the outcomes of any RTC depend on the theoretical definitions of disability adjusted life years (DALYs) and years of life lost (YLL). Here, DALY refers to the quantified health gaps as distinct to health to heal expectancies, and YLL refers to the years lost due to untimely death due to accidents (5).

Because children, pedestrians, and bicyclists often rely on the road for their daily activities, they are the most vulnerable and susceptible to traffic crashes (6). Each year, approximately 1.35 million people die from RTC, the leading cause of death in the 5-29 age group (7). In developing countries, safety measures to prevent accidents are almost entirely negotiable, resulting in pedestrians being involved in accidents more frequently than non-pedestrians (7, 8). This has led to a significant increase in accidents, especially among road users. This has ultimately had a negative impact on human, economic, and social costs within a nation and its communities (8).

In 2004, an estimation of road traffic collision showed that approximately 2.7 percent of total DALYs losses were due to RTCs, and this proportion is expected to increase to 4.9 percent by the year 2030 (9). RTC was reportedly accounted for 36% of the DALYs in 2017 (10). These data provide a clear explanation for the placement of road traffic accidents in the position of the third leading cause of global burden of disease worldwide. In low- and middle-income countries (LMICs), costs of 1 to 1.5 percent of GDP are required for road traffic crashes indicating that the health care budgets are particularly constrained in LMICs like Nepal (11). Some of the world’s most rugged and difficult mountain terrains, covering around three-fourths of the total geography, and so, having roads as most commuting modes of transportation, are furthering the RTCs in Nepal. However, evidence of the lost lives in such crashes and their economic cost is scant. Hence, the study aims to estimate the loss of DALYs and total cost associated with RTC.

## 2. Material and method

### 2.1. Study Setting

Kaski, a hilly district and the headquarters of Gandaki Province of Nepal, which contains the largest metropolitan (Pokhara, geographically) of the country, was selected as a study area. Surface transportation facilities, including feeder roads to national highways, district roads, and village roads, are increasing significantly, and district and rural roads are in poor condition and in need of regular maintenance (12). Nepal is basically a hilly country (nearly 83% geography is covered by hills and mountains), therefore, these conditions have also been regarded as direct or indirect reasons behind the yearly increase in road traffic crashes in Kaski district, and also, at national level.

### 2.2. Study Design and Sample Size

It was a retrospective, cross-sectional, institution-based survey. Taking reference of RTI prevalence from national level sample survey in 2014 (13); 2.67% (p), 5% (E), 2.5 (DEFF), and deduction with formula from the OpenEpi calculator (14) as [DEFF*Np(1-p)]/ [(d^2^/Z^2^_1-α/2_*(N-1)+p*(1-p)], taking hospitals as clusters and 10% non-response rate, we obtained a sample size of 110. We reached out to 116 RTC victims registered from one year to 45 days prior to the survey (conducted in November 2017), in five out of 16 hospitals (from emergency ward register records), traffic and police offices, and insurance office of Kaski district. Of 116 records, only 107 victims were eligible for inclusion (fig. 1).

**Figure 1.**
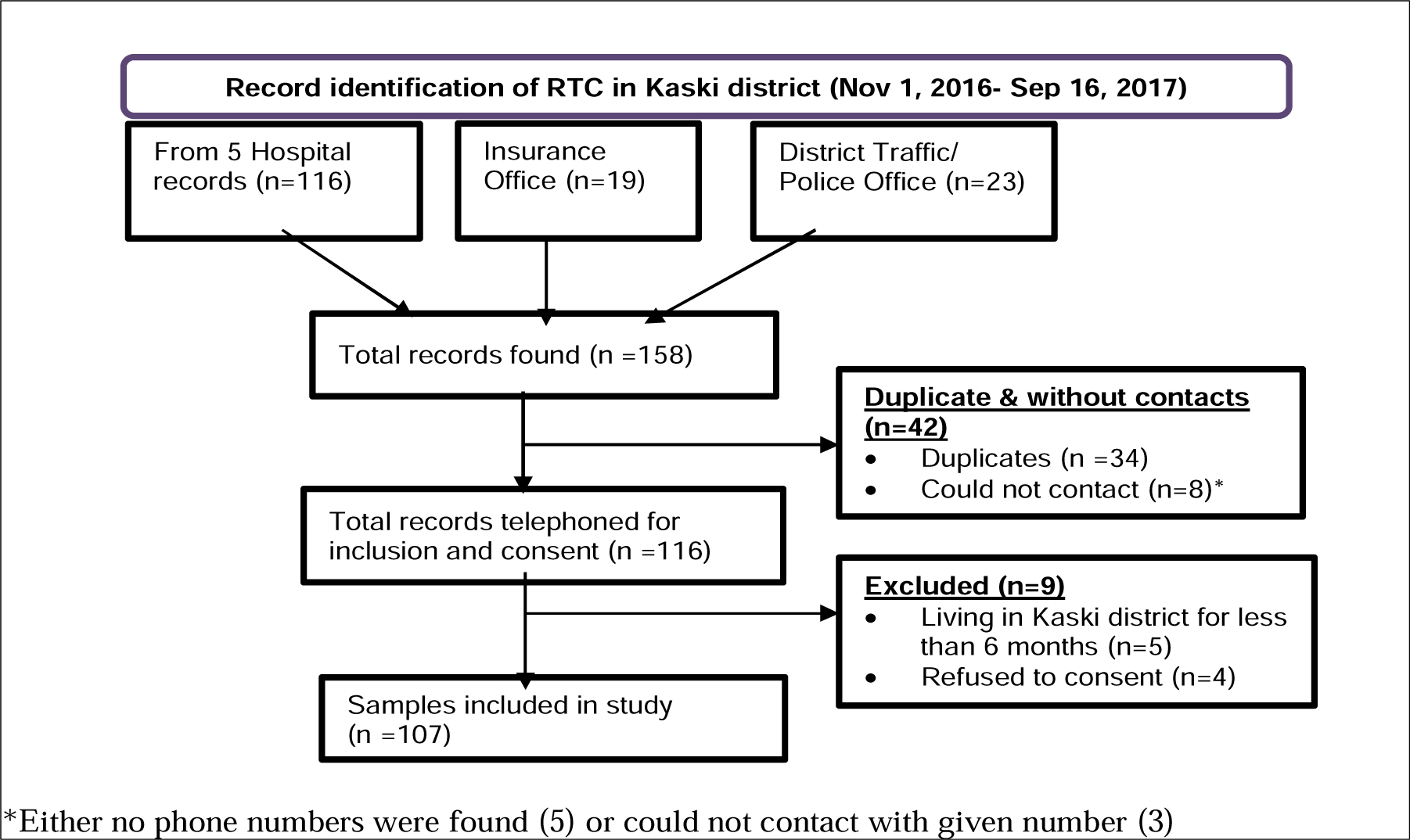
Sampling frame

**Fig. 2.**
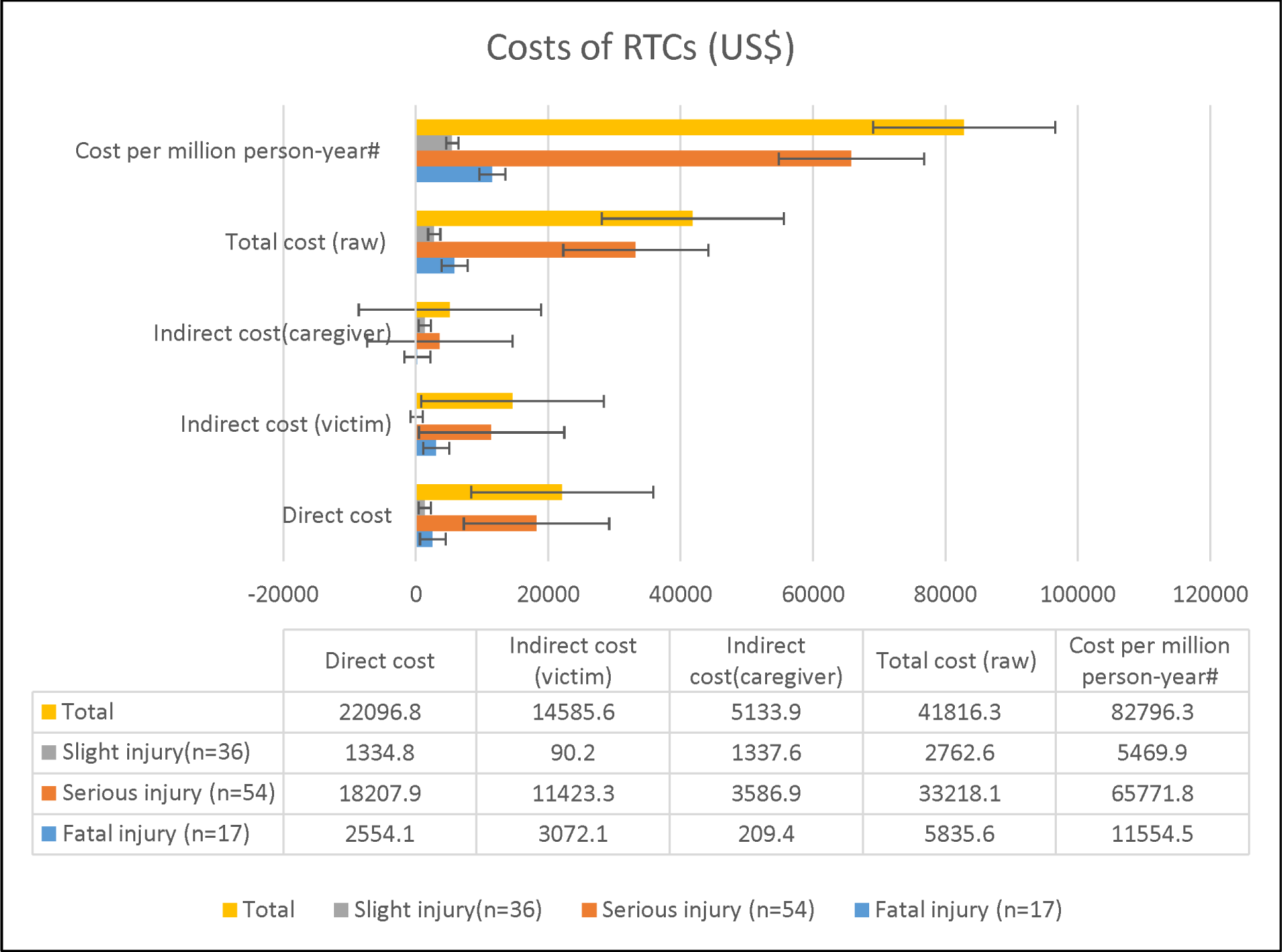
Costs of RTCs in US $ (US $1 =NRs 102.92 as per 2017, Jun 1) with standard error bars; ^#^Adjusted per million person-years (multiplied by factor 1.98 calculated from ((12/10.5)*(1000000/573025.2))

### 2.3. Data Collection, Ethics and Quality Assurance

In a predesigned and tested record format, we filled on the victims’ names, addresses, contact numbers and types of injury. With the help of the phone numbers provided in the documents, we contacted the victims or their family members for permission and date to interview. The pro forma was tested among 20 RTC victims of Syanja, a similar hilly and boundary shared with Kaski district, and then was used for the study individually at victim’s house or his/her convenient place. The pro forma consisted five sections-socio-demographic characteristics, economic status, information on accidents, direct and indirect costs accrued to victims and their caregivers and the information of disabilities-adjusted life years of the participants. After getting ethical clearance from Pokhara University Research Center (Ref no. 26/074/75), data were collected through face-to-face interview technique. In case of victim’s inability to respond or posthumous, we interviewed his/her nearest kin. Ethical norms regarding respondents’ dignity and confidentiality were maintained as per Helsinki and subsequent declarations. Responses regarding both costs-accrued to victims and the accidents were verified through prescription slips and medical bills and vouchers. In case of unrecorded costs and ambiguities, we further probed and assessed.

### 2.4. Statistical Analysis

Before being imported into the Statistical Package for Social Science (SPSS) and MS-Excel for further analysis, raw data were cleaned, coded, and entered into EpiData version 3.1. International Wealth Index (IWI) was used for economic status, in which 12 questions were asked and household level IWI scores were collated with MS-Excel and then in EpiData software. Cost calculation and converting it to a figure, and DALY’s estimation were also performed in MS-Excel.

### 2.5. Cost Calculation

Cost calculation was categorized as direct and indirect, and the calculation was done accordingly. Direct cost calculation accrued as the summation of all the costs that were paid for the medical treatment including all the bills starting from the registration charge to follow up. Indirect costs are not directly related to the cost of treatment and productivity loss, so, for the calculation of productivity loss, if any victim has died and fell into the category of economically active age group, average daily wage rate of a minimum of Nepalese rupee (NPR) 318 (US $ 3.1) (15), according to the Nepal Government, which was applied for the unemployed and whose monthly income was unknown. The indirect costs included were-transportation charges, bed charge, food charges and additional costs. The costs for both victims and caretakers were taken into account, and total indirect cost was calculated using the standard formula (16). The costs were asked and recorded in NPR and later transformed into US dollar as per exchange rate.

For

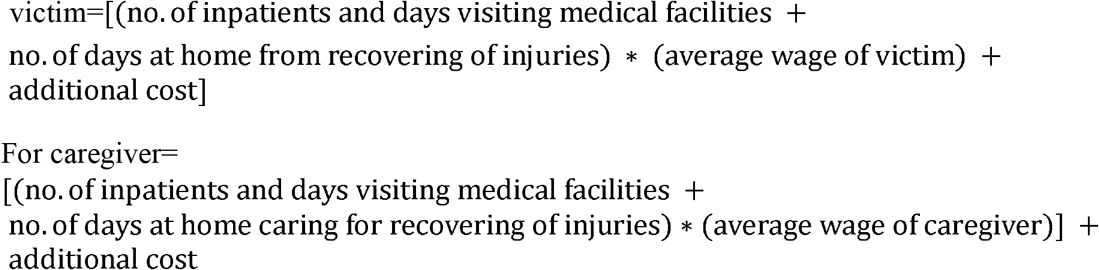

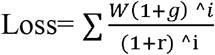, the formula, was used to measure the productivity loss where,

W denotes annual per capita gross domestic product, i.e., NPR 699.19 (US$ 1= NRs102.92), g, the annual economic growth rate, i.e., 0.041 (17), r, the discount rate, i. e., 0.03 (18), and, i, the average number of years lost due to fatal injuries (age of people died subtracted from standard life expectancy, i.e., 71 years).

### 2.6. Calculation of DALYs

Disability-adjusted life years are calculated with the sum of years of life lost and years of life lived with disability (YLDs), as used in 1990 and the 2004 global burden of disease (GBOD) studies (19), as follows:

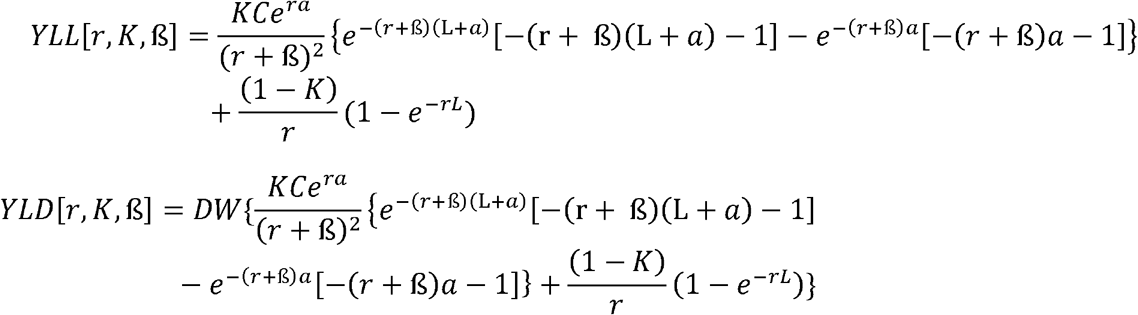

Here, the assumptions for discount rate (r) of 0.03, age-weighting modulation constant (K) of 1, parameter from the age-weighting function (β) of 0.04, and age-weighting correction constant (C) of 0.1658 were taken from the standard values given by the Global Burden of Disease Study (8, 20).

In the case of YLL, the age of death (in years) is denoted by “a” and standard life expectancy at the age of death (years) is denoted by “L”.

In the case of YLD, age of onset (years) is denoted by “a” and duration of disability (years) is denoted by “L”. DW (disability weight) was assigned based on severity weights ranging from 0 (representing perfect health) to 1 (representing death) as per protocol. Both YLL and YLD were measured for 107 victims for 10.5 months, and then later extrapolated to one year (with multiplying by 1.14, i.e., 12/10.5) and per million (with multiplying by 1.74, i.e., 1000000/573025.2).

## 3. Results

### 3.1. Socio-demographic Characteristics

Out of the total (n=107) victims, more than two-thirds (71%) belonged to the 15-44 year-age group. Similarly, the majority (60.7%) of the participants were male, and almost half (47%) were secondary graduate. A total of 59.8% were employed while the rest were unemployed. Sample population revealed significant differences by age group, ethnicity, education level, economic level, and occupation (p’s<.05) (table 1).

**Table 1.**
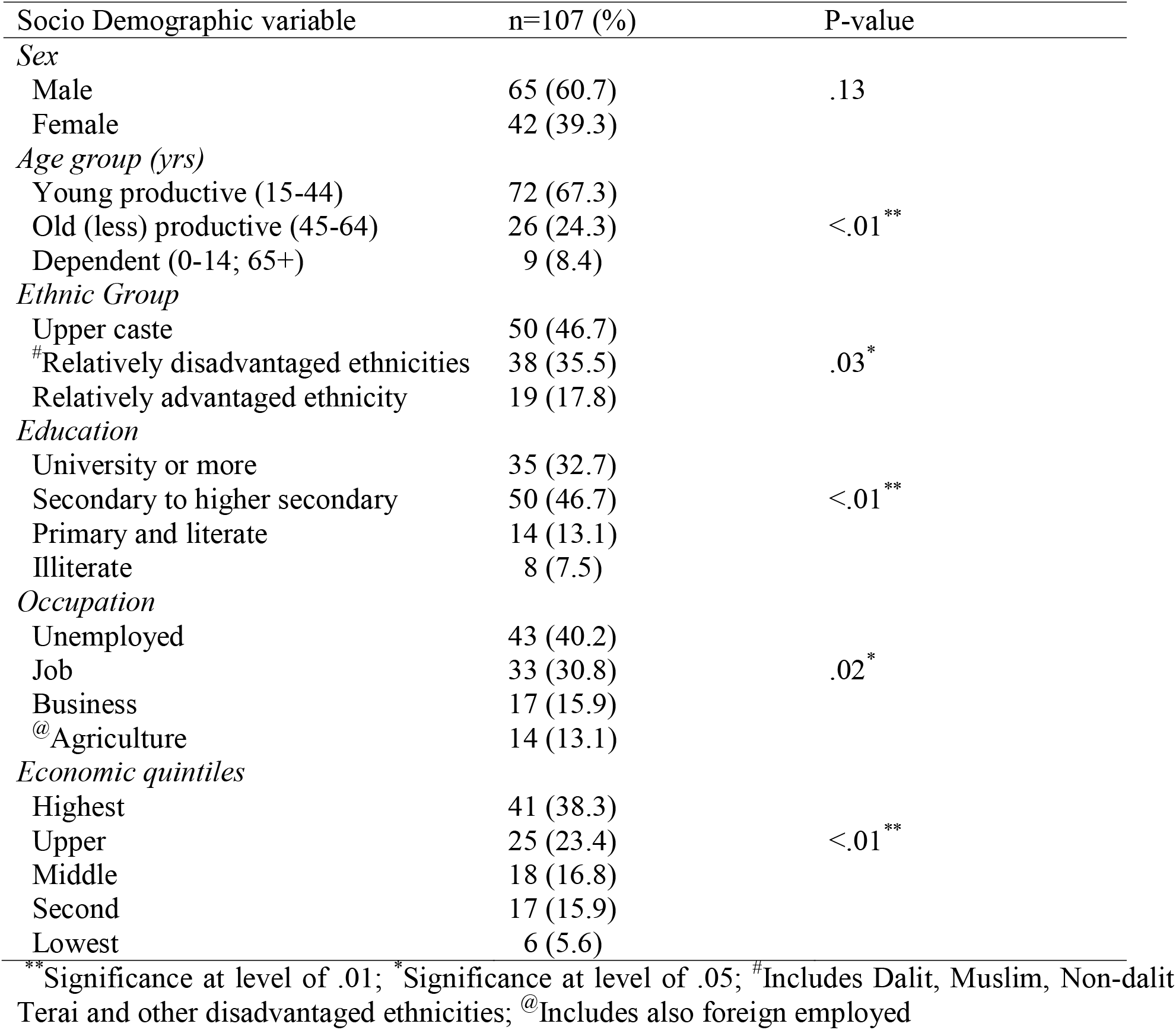
Socio-Demographic characteristics of respondent

### 3.2. Road Traffic Crash and its Various Dimension

Of 107 victims, more than four-fifths (82%) were injured and still suffering from different types of disabilities during the survey period, and the rest (18%) had lost their lives. Adjusted to per million person-years, deaths due to RTC were 37.7. Serious injury was high (p<.01) with 50% compared to slight injury (33.6%) and fatal injury (15.9%). According to the findings, motorcyclists were the most affected (frequency, 49%, p<.01), followed by bus and truck drivers. There are several reasons behind the accident-overtake/ over speed (27.1%) stood the highest in proportion, followed by carelessness of the driver/ pedestrian (25.2%) (table 2).

**Table 2.**
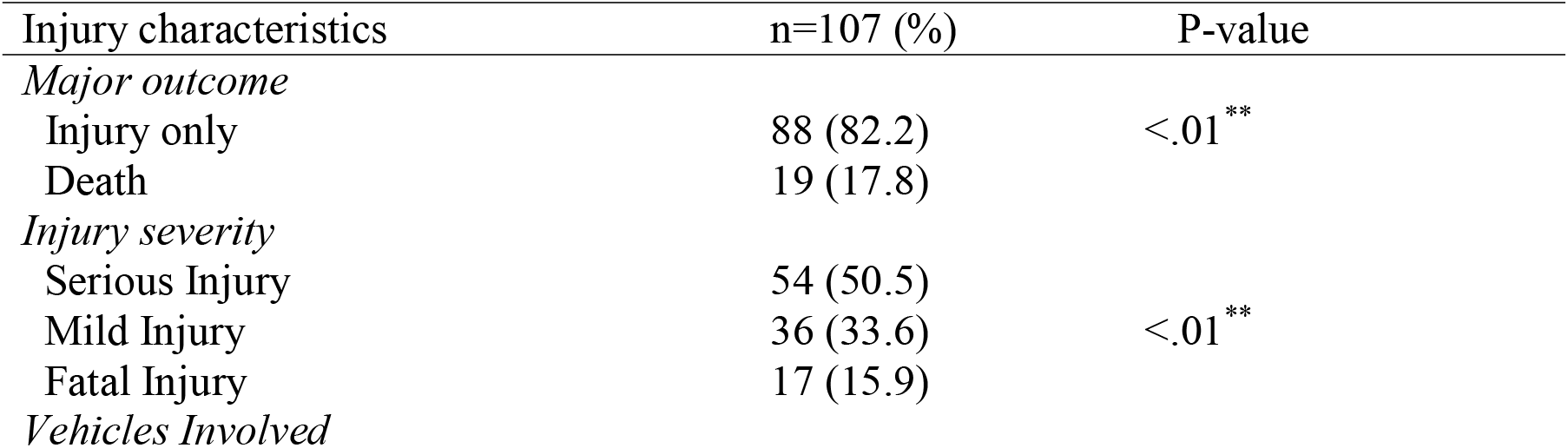
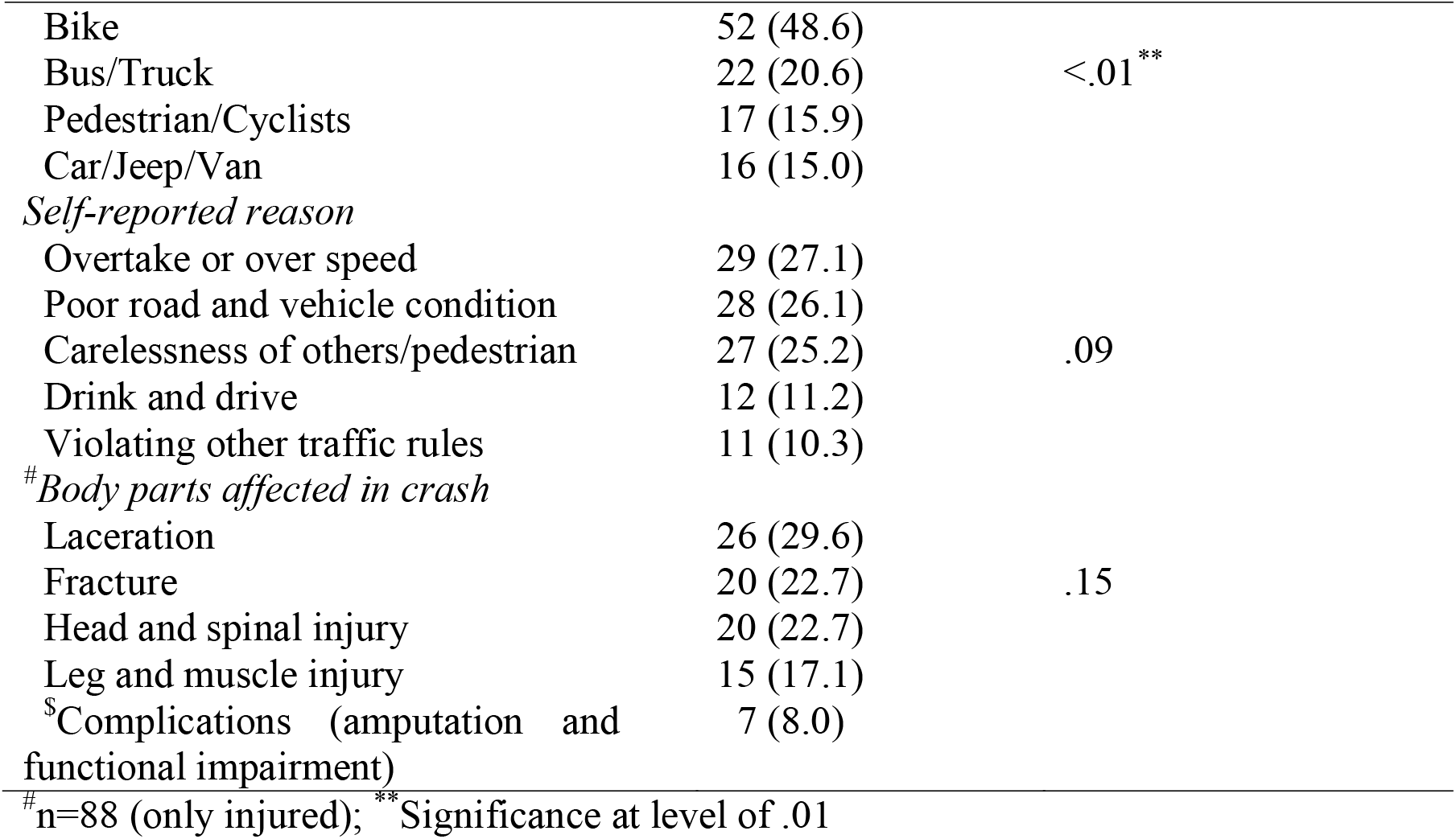
Frequency distribution of injuries and vehicle involvement of RTC

| Injury characteristics | n=107 (%) | P-value |
| --- | --- | --- |
| *Major outcome* |  |  |
| Injury only | 88 (82.2) | <.01** |
| Death | 19 (17.8) |  |
| *Injury severity* |  |  |
| Serious Injury | 54 (50.5) | <.01** |
| Mild Injury | 36 (33.6) |  |
| Fatal Injury | 17 (15.9) |  |
| *Vehicles Involved* |  |  |
| Bike | 52 (48.6) |  |
| Bus/Truck | 22 (20.6) | <.01 ** |
| Pedestrian/Cyclists | 17 (15.9) |  |
| Car/Jeep/Van | 16 (15.0) |  |
| *Self-reported reason* |  |  |
| Overtake or over speed | 29 (27.1) |  |
| Poor road and vehicle condition | 28 (26.1) |  |
| Carelessness of others/pedestrian | 27 (25.2) | .09 |
| Drink and drive | 12 (11.2) |  |
| Violating other traffic rules | 11 (10.3) |  |
| *#Body parts affected in crash* |  |  |
| Laceration | 26 (29.6) |  |
| Fracture | 20 (22.7) | .15 |
| Head and spinal injury | 20 (22.7) |  |
| Leg and muscle injury | 15 (17.1) |  |
| *\$Complications (amputation and functional impairment)* | 7 (8.0) | |

#n=88 (only injured); \*\* Significance at level of .01

### 3.3. The Cost of Road Traffic Crashes

The total cost of NPR 4303724.3 (US $ 41844.7) was split into two parts: the direct cost (NPR 2274196.0 (US $ 22096.7)) was slightly higher than the indirect cost. However, it was the inverse in the cases of less severe and fatal injuries. The currency conversion was done as per exchange rate of Central Bank of Nepal (Nepal Rastra Bank) dated December 15, 2017 (15) (figure 1). The dissimilar standard error bars show the unequal variances between the costs.

### 3.4. Total DALYs Lost

Total numbers of YLL and YLD totals were 1011.8 and 8.8 per million person-years, respectively. Almost all the proportion of DALYs (99.0%) was shared by YLL (table 3).

**Table 3.** Total DALYs Lost during the period of 1^st^ Nov, 2016-15^th^ Sep, 2017

| SN | Indicator/Duration | Burden of RTC |  |  |
| --- | --- | --- | --- | --- |
|  |  | Year of life lost (YLL) | Year Lived with Disability (YLD) | DALYs (YLL+YLD) |
| 1 | Raw scores from survey (10.5 months) | 508.68 | 4.43 | 513.11 |
| 2 | *#*Adjusted (1 year) | 579.80 | 5.05 | 584.85 |
| 3 | *\$*Per million person-year (2017) | 1011.82 | 8.81 | 1020.63 |

*#*Survey raw score, multiplied by 1.14 (to convert into 12 months);
*\$*Population projection of 2017 (573025.2) of Kaski district based on Census 2011 (492,098 and PGR=2.57)<sup>[38]</sup>

### 3.6. Correlation between the Cost and DALYs

The indirect cost for the victim (r=0.33, p<.001) and total cost (r=0.2, p=.039) were moderately and lowly correlated with DALYs, respectively, whereas with DALYs, the indirect cost for the caregiver (r=-0.044, p>.05) and the direct cost (r=0.159, p>.05) were statistically non-significant (table 4).

**Table 4.** Correlation between Cost and DALYs

| Variables | DALYs |  |
| --- | --- | --- |
|  | Pearson r | P value |
| Indirect cost of victim | 0.33 | <.01 ** |
| Indirect cost of care giver | -0.04 | .65 |
| direct cost | 0.16 | .10 |
| Total cost | 0.20 | .04* |

\*Statistically significant at p<0.05; \*\* statistically significant at p<0.01

## 4. Discussion

Calculations were based on total medical expenses, productivity loss, lost income during the accident and survey period, and different indirect costs. The study findings reflect the portentous situation of the RTC and the victims in Kaski district. The pronest groups, costs, types of injuries, DALYs, their impacts, and the associated factors are discussed.

The majority of RTC victims were male and employed (both more than 60%, of which 30% were in service jobs). Most victims felled in the 25-44 year-group (37.4%), had a university degree (33%), and belonged to the highest economic quintile (38%). A similar study among brain injury victims among motorcyclists, carried out in Nigeria, showed that nearly three-fourths (73.96%) felled in the 20-50 year-group (21). Despite similar topography but different elevations from sea level, as compared to 121 deaths in Sana’a city in Yemen, ours had a much lower (37.7) mortalities per million person-years (22). This may lead to further research in Kaski, and country-wide, focusing on mass transport, speed limits, and gross anatomy of the roads like straightness, levels, and grades. According to the Sana’a city study, more than half of RTCs occurred in straight roads (22), while another study conducted in Iran found that more than three-fourths of RTCs (77.6%) occurred on straight and level roads, followed by 13.5% on curve and level, and then negligible on curve and grade (5.3%) and straight and grade (3.6%) roads (23). These factors should be explored in the context of Kaski and further research. Another major impact is on economies, as most victims are the breadwinners, who generate the economic returns for the family, community, and nation. Studies conducted in the capital metropolitan area (8) and in eastern Nepal (24) have shown that the proportion of men is even higher. These findings are also in line with those of other countries, such as Valencia, Spain (25), Pakistan (26), Haryana, India (27) and Tanzania (28). Furthermore, it was revealed that nearly half of the accidents involved motorcycles and one fifth involved buses and trucks. Similar results were also found in studies conducted in other cities in Nepal(8, 29) and in Kenya(30). In addition, pedestrians were also seen as one of the affected groups (29) due to the poor knowledge of road safety. High speed and overtaking, followed by carelessness of riders or pedestrians, road conditions, drinking and driving, violation of traffic rules and vehicle conditions, are some of the reasons found in the RTI studies carried out by Thapa (29) and Sapkota et al.(8). The observed risk of males to be victimized may be due to their risk-taking behavior fueled by alcohol consumption, mobility in nature, and engagement in outdoor activities more than their female counterparts, resulting in a heightened risk of accidents. This is further supported by the findings of the study in Kathmandu by Sapkota et al. (8), in Ghana (31), WHO global report (30) and Vietnam (31). The main reasons for overtaking and negligence from our study were consistent with driver negligence as the main cause, followed by high speed and overtaking, are revealed in the WHO report (16) and studies carried out in other cities of the country (8, 29).

The male workers being victimized to a greater extent in our study reflects the higher proportion of men in the sample as well as the ratio of male to female workers in Nepal.

The finding that more than a half of the victims were found with severe injuries is also consistent with the study conducted among the victims of motorbike accidents in Kathmandu, Nepal (8, 29, 32). The mortality rate following RTC in the present study of 16% is also consistent with that conducted in other developing countries, where it ranged from 10-30% (33, 34). Similar to our findings, a community based study from Nepal showed that lacerations were the most common type of injury followed by fractures (8).

DALYs, with indirect cost of victim (r=0.33; p<0.001), and with total cost (r=0.19, p=0.039) had low positive correlations, respectively. Descriptive statistics showed that indirect costs were higher than direct costs in minor and fatal injuries, whereas it was the inverse for serious injuries (table 3). In line with slight and fatal injuries, higher indirect costs than direct costs were noted in the study of motorbike accidents in Kathmandu (8). Because of the productivity loss of the victims and the caregivers, calculations for indirect costs might have shown the higher values. Variations in costs from one to another country may be found due to the variations in health-care delivery systems, insurance provision and patterns, service costs, driving culture and traffic rules, and many more.

The majority of the victims in these studies were the household heads and breadwinners of the families, and their deaths and injuries could have potentially long-term implications for the financial sustainability and social well-being (8, 30). Moreover, the out-of-pocket expenditure (OOP) incurred for the treatment of the victim leads the family to further impoverishment (34). So, the economic impacts of the injuries, mainly the RTIs, are both high in OOP and in productivity loss (8, 35, 36).

The total cost and the DALYs associated with RTC in Kaski district in 2017 were around US $ 82800 and 1021 per million person-years, respectively, as of the projected population of the country in 2017 (37, 38). A similar study carried out in Kathmandu valley, the capital city, and then extrapolated at a national level, found a much lower cost, i.e., US $ 5993 per million person-years of population (39). This larger variance indicates that Pokhara loses a greater proportion of the cost associated with RTC when compared at the national level. However, the estimated costs from the societal perspectives (after calculating incidence-based cost-of-illness) at the national level in 2020 showed a quite higher loss of the cost, that is US $ 463,794 per million population (39). These significantly higher costs compared to our study are due to the costs added for grief, pain and suffering, and other production costs (39). However, the proportion of direct costs in our study was significantly higher (52.8%) than in a study (10.6%) to estimate the costs from societal perspectives by Banstola et al. (39). A burden of injuries study from secondary data of the GBOD study 2016 showed a DALY loss of 14,052 per million person-years from ‘transport injuries’ of which 90% (around 12647) is from RTCs (40). It showed that the national average of DALYs lost is quite higher than the findings of current study (around 1018 per million person-years), which may be due to the lower coverage in the current study.

### 5. Limitation

The study has many limitations. Firstly, the limited sample of 107 RTC victims, mainly from Pokhara valley and the surroundings (as some hospitals refused to give permissions), may be less representative. Also, from our sample, a minimal YLL value was obtained as compared to YLD, which should also be taken cautiously. Second, victims who were tranferred outside Pokhara for treatment (possibly to Kathmandu, where the national trauma center and other tertiary level hospitals are located, as well as to India) could not be covered. Thirdly, many minor and some moderate cases that were not taken to hospitals have also been missed. Fourthly, the study could not cover the cost of vehicle and general damage, future medical expenses, and the cost of suffering and other societal values attributable to the crash, which have not been calculated, and finally, the cost attributable to co-morbid conditions of the victims has not been adjusted. Despite these limitations, the primary data-based cost and DALYs of RTC are scarce and so, recommended to apply with caution.

## 6. Conclusion

Per million person-years cost and DALYs of RTC were approximately $82,800 and 1021, respectively. The young unemployed motorcyclists were at risk and serious injury is probably a most frequent outcome. The victim’s indirect cost increased with DALYS, suggesting a policy implication, but may need a further reaffirmation.

## Data Availability

All data produced in the present study are available upon reasonable request to the authors.

## Conflicts of Interest

We declare that we have not any conflicts.

## Funding

We did not receive any funding for this study.

## Authors’ Contribution

SB and CA conceptualized the study. SB and DK collected the data. SB and CA analysed the data. DK, CA, SB, BG, VPS and CLR contributed to the first and subsequent drafts. CA further reviewed and edited the reviewer’s comments. All the authors contributed and reviewed the final manuscript, and approved for submission.

## Acknowledgement

We duly acknowledge Mr. Chandra Prasad Khanal, IELTS Instructor, Valley International Consultancy Pvt. Ltd., Kathmandu, Nepal, for language editing.

## Notes

### Competing Interest Statement

The authors have declared no competing interest.

### Funding Statement

This study did not receive any funding.

### Author Declarations

Ethical approval was granted from Institutional Review Committee (IRC) of Pokhara University Research Center (PURC), Kaski, Nepal.

### Summary of Updates

Results section mainly, has been updated to show the p-values of difference in frequencies of socio-demographic factors (table 1), and injury related factors (table 2); and revised in text of results, discussion and abstract, including the conclusions.

